# Replication of recipient genetic variants associated with survival after lung transplantation

**DOI:** 10.64898/2026.11.17.26368313

**Authors:** Kati Hyvärinen, Jukka Partanen, FinnGen, Jouni Lauronen, Jarmo Ritari

## Abstract

**Background:** Long-term survival after lung transplantation (LTx) remains limited, largely because of chronic lung allograft dysfunction and other post-transplant complications. Genetic studies have suggested recipient single-nucleotide polymorphisms (SNPs) to associate with LTx outcomes, but none has been confirmed in independent cohorts.

**Methods:** We evaluated 29 previously published candidate SNPs associated with LTx complications in 201 LTx recipients from FinnGen. Time-to-event analyses were performed for death after transplantation, with censoring at end of follow-up. Each SNP was tested under additive, dominant, and recessive genetic models using Cox proportional hazards regression adjusted for sampling age, sampling year, sex, and the first ten genetic principal components. Replication was defined as nominal association with survival at p < 0.05, surpassing minimum detectable effect at 80% power and concordant direction of effect with the original publication.

**Results:** We found two independent replicated signals: rs4526713-T and rs4149117-T. In multivariable analysis, rs4526713-T and rs4149117-T were associated with increased mortality risk under an additive and dominant models, respectively. A two-SNP score showed a stronger association with mortality than any individual SNP, suggesting an aggregate contribution of replicated genetic risk alleles to post-transplant survival.

**Conclusions:** Our findings support the contribution of host genetic variation to long-term LTx outcomes. However, modest cohort size and use of all-cause mortality rather than allograft dysfunction-specific outcomes require cautious interpretation. Larger donor–recipient genomic studies with harmonized transplant phenotypes are needed before clinical implementation.

## Introduction

Lung transplantation is an established treatment for selected patients with end-stage lung disease, but long-term outcomes remain poorer than for many other solid-organ transplants (Michel et al., 2025). Chronic lung allograft dysfunction (CLAD) is a major barrier to durable graft function and survival, and is characterized by progressive, often irreversible decline in respiratory function. Recent European data also emphasize substantial variability in CLAD diagnostic work-up and management, reflecting both biological complexity and incomplete standardization across transplant centers (Gottlieb et al., 2025). Advanced CLAD is associated with considerable morbidity and mortality. In a recent European cohort of patients with advanced CLAD, two-year survival was only 25%, underscoring the clinical importance of identifying patients at increased risk (Kovacs et al., 2025).

The pathogenesis of adverse outcomes after lung transplantation is multifactorial. Established contributors include donor and recipient characteristics, primary graft dysfunction, acute cellular and antibody-mediated rejection, infection, alloimmune injury, HLA mismatch, immunosuppressive exposure, and non-immune injury (Napoli et al., 2024). Increasing evidence suggests that inherited recipient genetic variation may also modify risk, either through immune regulation, epithelial injury responses, complement activation, drug metabolism, or susceptibility to rejection and CLAD. A 2015 review concluded that numerous polymorphisms had been associated with lung-transplant complications but also emphasized that many findings lacked independent replication and that larger multicenter studies were needed (Ruttens et al., 2015).

The GenCOLT biobank, based on the multicenter COLT cohort, combined donor and recipient DNA, genome-wide genotyping, HLA data, and clinical phenotyping for 387 LTx pairs, including more than 7.3 million SNP and HLA genotypes after quality control (Brocard et al., 2025). Recently, a genome-wide association study in LTx reported two recipient loci associated with CLAD and confirmed the deleterious contribution of HLA allelic and epitopic mismatches, particularly class I HLA mismatches (Brocard et al., 2026). These developments highlight the potential value of genomic data but also illustrate that robust inference requires replication across independent transplant cohorts.

In this study, we sought to replicate previously reported recipient SNP associations with LTx complications in an independent Finnish biobank cohort. Because detailed CLAD phenotyping was not available, we used survival after LTx as a clinically meaningful endpoint. We further assessed whether replicated variants exerted independent effects and whether an aggregate SNP score improved risk stratification compared with individual variants.

## Materials and Methods

### Study population and lung transplantation cohort definition

FinnGen is a public-private partnership collecting and analyzing genotype and medical phenotype data from half a million Finns (https://www.finngen.fi/en) (Kurki et al., 2023). Contributing institutional members of FinnGen and their affiliations are available in Supplementary Table 1. Lung transplant recipients were identified from FinnGen release R14 longitudinal phenotype data using the operation code GDG10 corresponding with transplantation of both lungs. Individuals with this code were extracted from the detailed longitudinal phenotype file, yielding 218 LTx records.

FinnGen microarray-based genotyping data were called and filtered for variant and sample quality, followed by phasing and imputation as described previously (Kurki et al., 2023). Detailed information can also be found at https://docs.finngen.fi/. Genotype data for the LTx recipients were extracted from FinnGen R14 PLINK genotype files using PLINK2 v2.00a6LM (Chang et al., 2015), restricted to identified LTx recipients and to the selected candidate SNPs. Extracted genotypes were converted to additive allele dosages using PLINK2’s --recode A option.

The extracted longitudinal phenotype data for the LTx recipients were then linked with FinnGen R14 covariate data and SNP allele dosage data. After merging transplantation records, covariates, and candidate SNP data, the final analytical dataset contained 213 observations corresponding to 201 unique individuals. Follow-up time was calculated as the difference between age at death or end of follow-up and age at lung transplantation.

Pre-transplant comorbidities were derived from FinnGen ICD-10 based longitudinal endpoint data by extracting all endpoint records occurring before lung transplantation. The following pre-transplant diagnoses were selected and summarized: chronic obstructive pulmonary disease (COPD), interstitial lung disease (ILD), asthma, cystic fibrosis, respiratory diseases principally affecting the interstitium with inflammatory bowel disease (IBD) comorbidities, pulmonary embolism, idiopathic pulmonary fibrosis, and diabetes. Baseline characteristics were summarized by sex using R package gtsummary v2.5.1 (Sjoberg et al., 2021). Study cohort characteristics are given in Table 1.

**Table 1.**
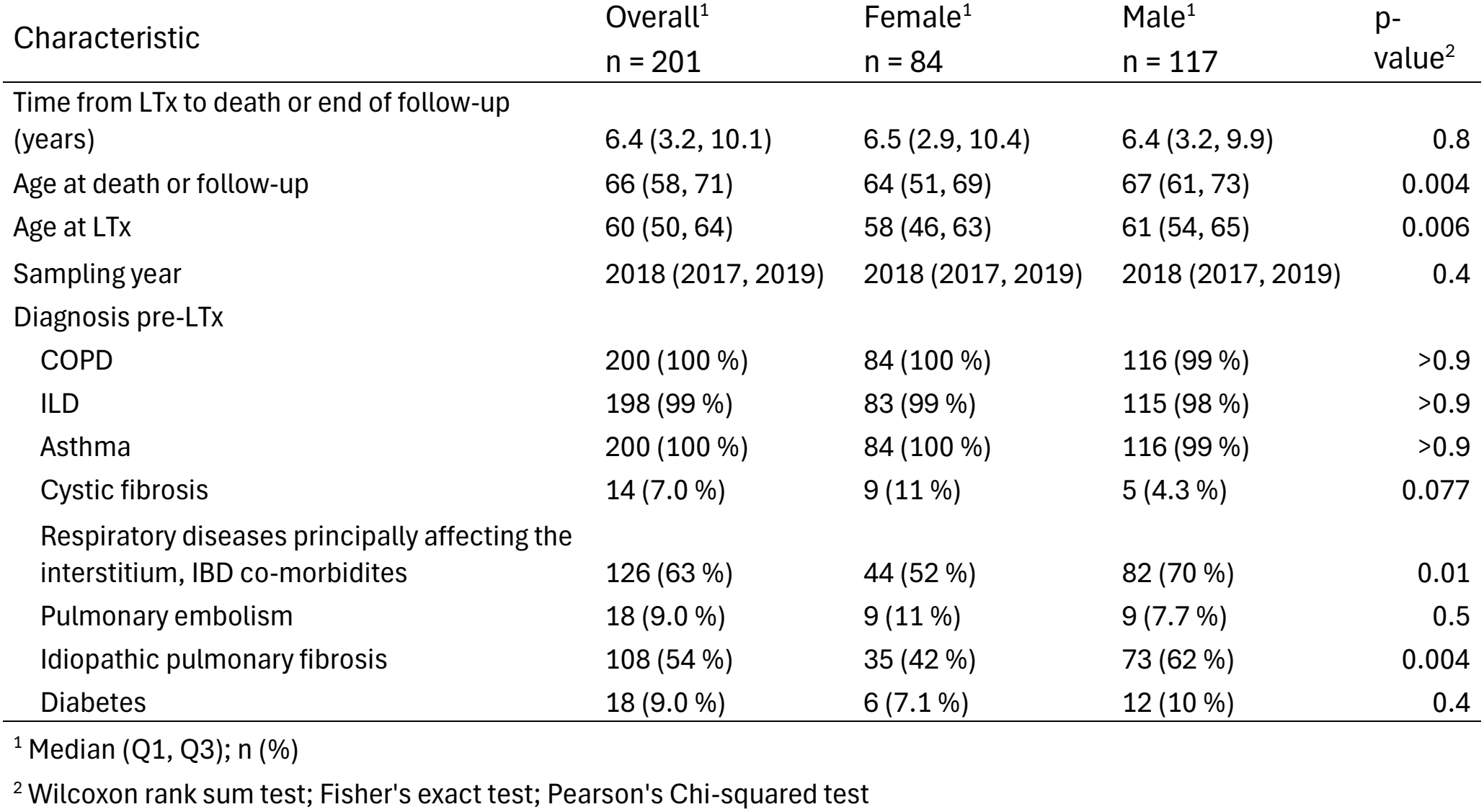
Study cohort characteristics.

### Ethical permits

Study subjects in FinnGen provided informed consent for biobank research, based on the Finnish Biobank Act. Alternatively, separate research cohorts, collected prior the Finnish Biobank Act came into effect (in September 2013) and start of FinnGen (August 2017), were collected based on study-specific consents and later transferred to the Finnish biobanks after approval by Fimea (Finnish Medicines Agency), the National Supervisory Authority for Welfare and Health. Recruitment protocols followed the biobank protocols approved by Fimea. The Coordinating Ethics Committee of the Hospital District of Helsinki and Uusimaa (HUS) statement number for the FinnGen study is Nr HUS/990/2017.

The Coordinating Ethics Committee of the Hospital District of Helsinki and Uusimaa (HUS) statement number for the FinnGen study is Nr HUS/990/2017. All FinnGen approval codes are available as Supplementary Data.

### Candidate SNP selection

We curated 29 candidate SNPs previously reported in association with LTx complications, including primary graft dysfunction, acute rejection, chronic rejection/CLAD, infection, pharmacogenomic response, or mortality. The variants were collected from articles identified based on PubMed search using search terms “lung transplantation snp” and “lung transplantation gwas”. We curated the search results manually, accepting studies that reported a statistically significant genetic association in recipients of LTx. Candidate variants and their source studies are listed in Table 2.

**Table 2.**
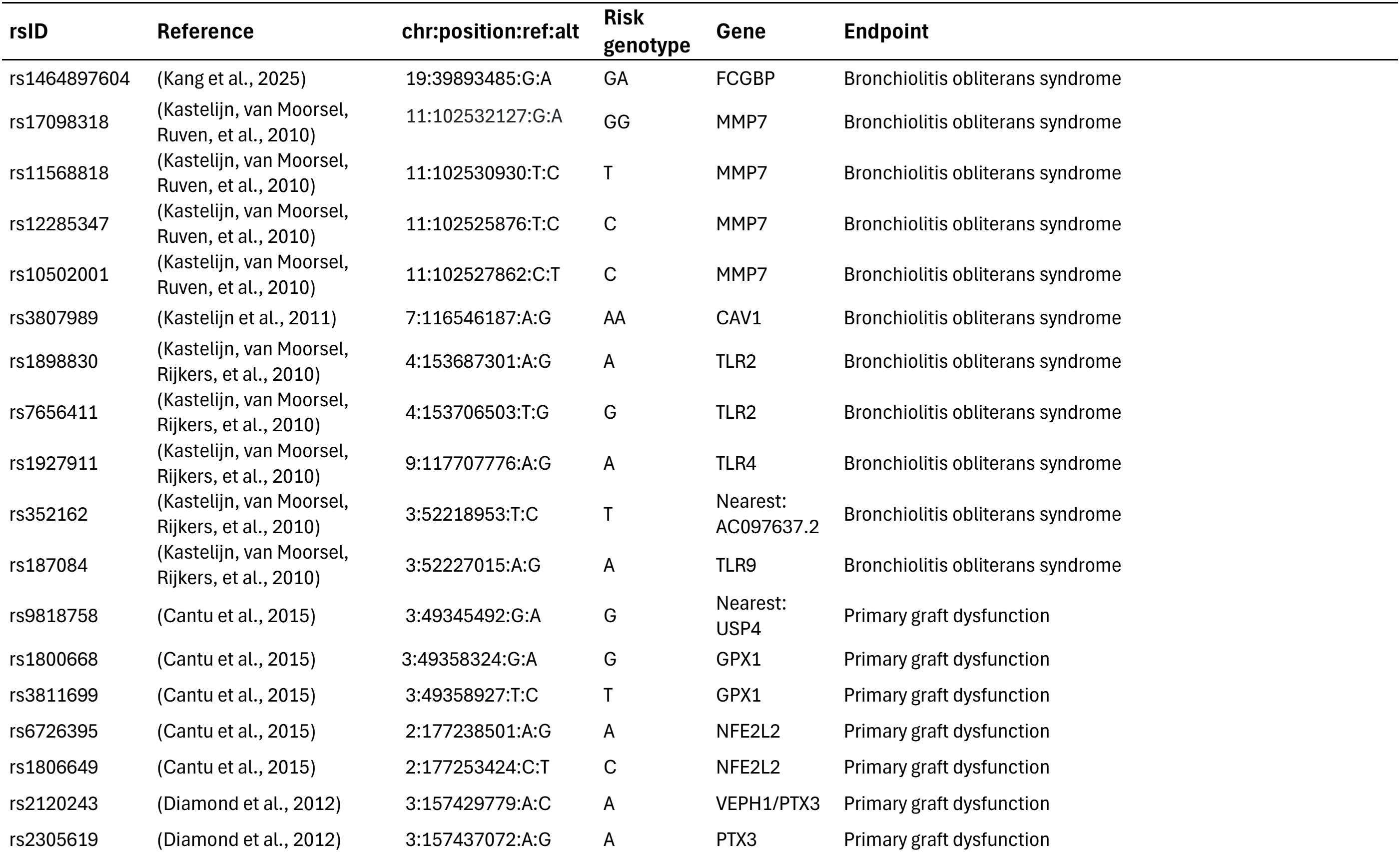
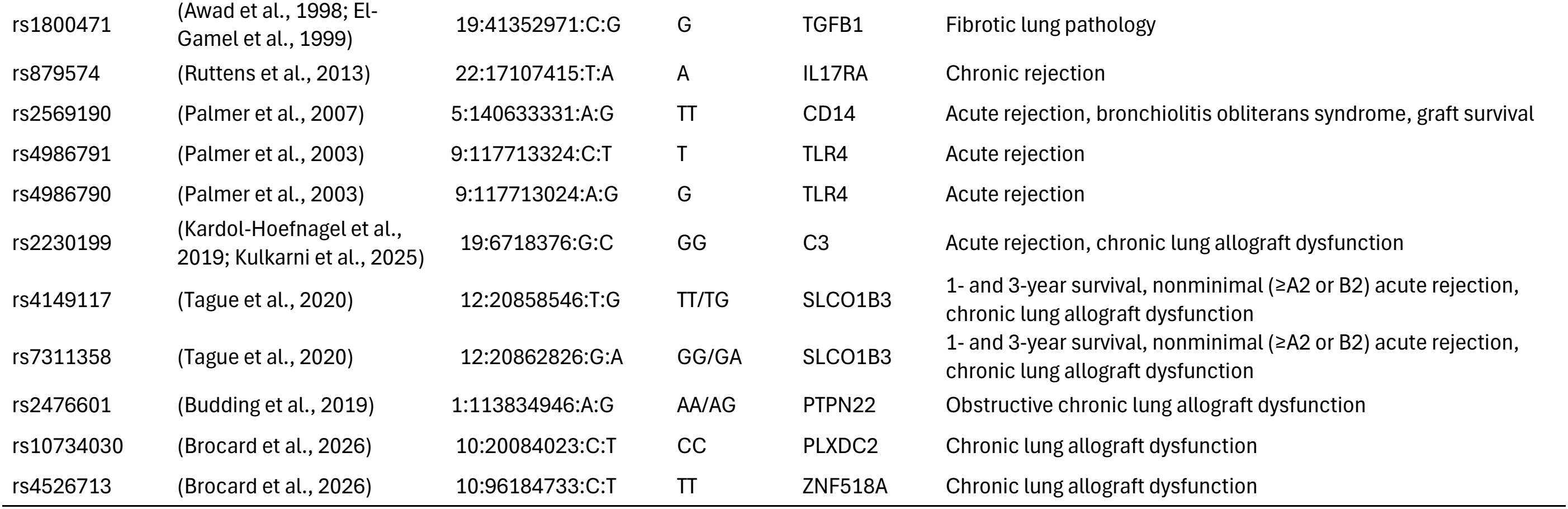
Candidate genetic variants in LTx survival curated from literature.

### Outcome

The primary outcome was time from lung transplantation to death. Patients alive at the end of follow-up were censored. Because outcome-specific transplant phenotypes such as CLAD, acute rejection, infection, and primary graft dysfunction were not available in the present analysis, all-cause mortality was used as an integrated clinical endpoint.

### Genetic models and association testing

Associations between candidate SNPs and post-transplant survival were evaluated using Cox proportional hazards regression. The outcome was time from lung transplantation to death or end of follow-up, with death status as the event indicator. Each SNP was tested individually in multivariable Cox models adjusted for the selected clinical and genetic covariates: sampling age, sampling year, sex, and the first ten genetic principal components. Three inheritance models were evaluated for each SNP: additive, recessive, and dominant. Additive models used allele dosage directly; recessive models compared homozygous risk-allele carriers with all other genotypes; dominant models compared carriers of at least one risk allele with non-carriers. Model estimates were converted to hazard ratios (HR) by exponentiation and reported with corresponding standard errors, confidence intervals, and p-values (Table S2). For each SNP, the model with the smallest p-value among the three inheritance assumptions was selected for replication assessment.

### Replication criteria

A candidate SNP was considered successfully replicated if it fulfilled three criteria: 1) nominal multivariable coxph association with survival at p < 0.05; 2) surpassing the minimum detectable effect size (in either direction) at 80% statistical power given p < 0.05, sample size of 201, event rate of 24.88% and SNP-specific minor allele frequency; 3) concordant direction of effect for the risk allele or risk genotype with the original report. The minimum detectable effect size for coxph hazard ratios (MDHR) was computed using the Schoenfeld approximation (Schoenfeld, 1983).

### Joint model and SNP score

The two replicated SNPs were included into a multivariable coxph model together with the covariates described above to assess whether each variant contributed independently to survival (Table S3). To model the combined effect of the two replicated SNPs, we summed them into a genetic score. The score was calculated as the average of dosages of the two genetic variables given their inheritance model, i.e., additive for rs4526713-T and dominant for rs4149117-T. As a result, the score had four possible values according to genetic risk: 0, 0.5, 1, and 1.5. The score was tested as a continuous predictor in a multivariable coxph model adjusted for the same covariates as described above (Table S3). For visualization, patients with a score of 0 or 1.5 were selected as low- and high-risk extremes.

### Kaplan–Meier visualization

Survival curves were generated and visualized using R package ggsurvfit v1.2.0 (Sjoberg et al., 2026). Curves were plotted for genotype-defined groups for the two SNPs and for high versus low SNP-score groups. For rs4149117, individuals were grouped as TT/GT versus GG; and for rs4526713, homozygous genotype groups were contrasted, excluding heterozygotes from the visualization for clarity. For the SNP score, individuals in the top of the score distribution were compared with those in the bottom. Survival plots included confidence intervals and annotations showing hazard ratios, 95% confidence intervals, and p-values from the corresponding multivariable coxph models.

### Protein QTL analysis

To investigate potential functional consequences of the top lung transplantation survival-associated SNPs, protein quantitative trait locus associations were queried from pan-asset pQTL meta-analysis summary statistics combining UK Biobank Pharma Proteomics Project (PPP) and FinnGen Olink proteomics data (n_total_ = 48,703). A BED-style region file was created for the two top SNP positions: chr10:96184733 and chr12:20858546. For each protein-level summary statistics file, tabix v1.16 was used to extract association results overlapping these SNP positions. Extracted results were combined across proteins, and p-values from the inverse-variance meta-analysis were adjusted using the Benjamini–Yekutieli false discovery rate method (Table S4).

### AI-use

During manuscript preparation, Microsoft Copilot (accessed October 2026) was used for English-language editing and improvement of readability. No AI tool was used for study design, data analysis, interpretation of results, or generation of scientific conclusions. All outputs were critically reviewed by the authors, who take full responsibility for the final content.

## Results

### Candidate variant replication

Among the 29 candidate SNPs (Table 2), three fulfilled the replication criteria (Figure 1). Two variants were located within 4.3 kB of each other and yielded effectively identical results (rs4149117 and rs7311358), suggesting redundancy due to linkage disequilibrium or equivalent genotype information in this cohort. Therefore, one representative variant was retained (rs4149117), leaving two potentially independent replicated signals for downstream analyses. The coxph summary statistics for all tested SNPs under the three genetic models are given Table S2.

**Figure 1.**
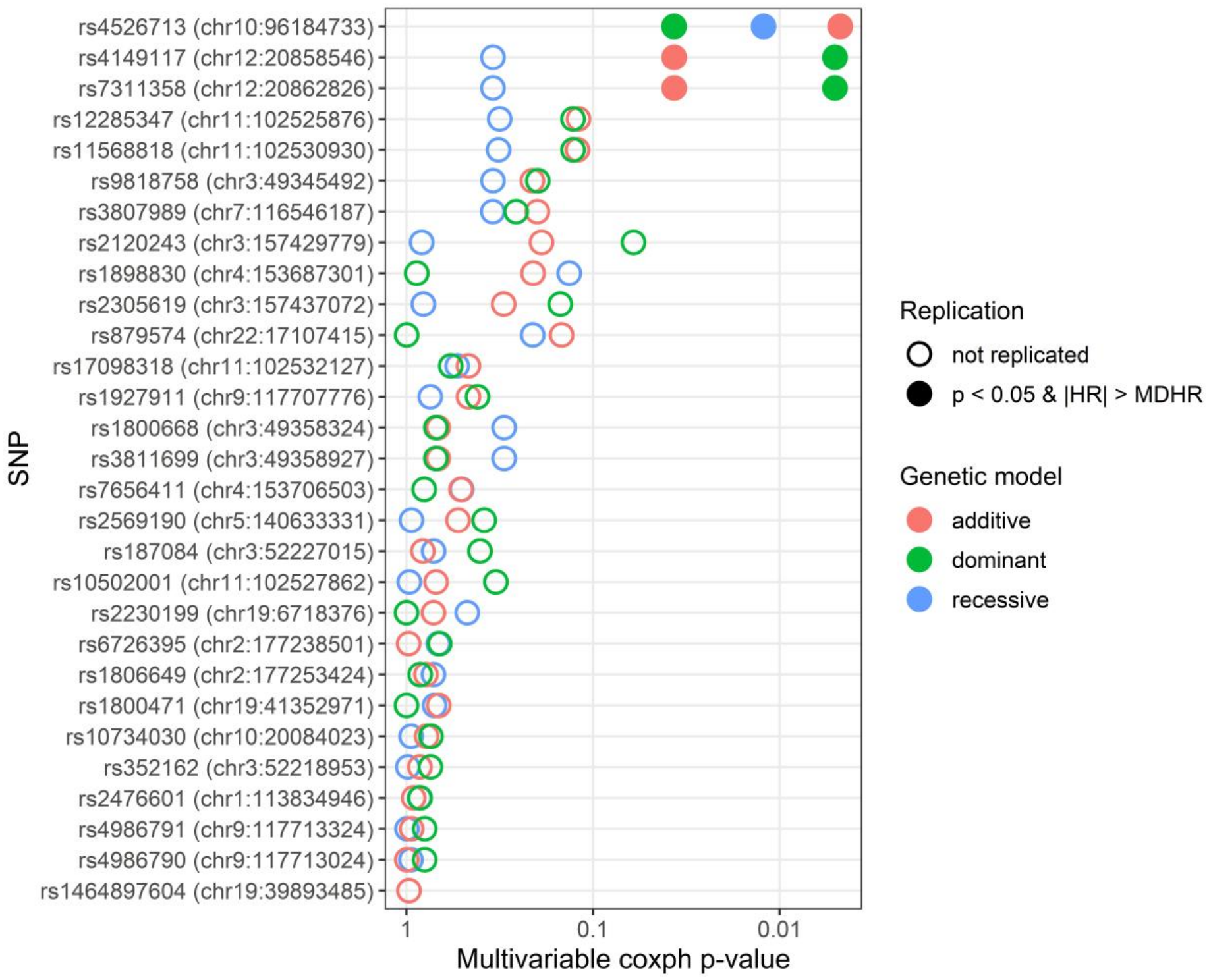
Replication test of candidate SNPs for survival time after LTx under three genetic models using multivariable Cox proportional hazards model (coxph). Model covariates were sampling age, sampling year, sex, and the first ten genetic principal components. MDHR; minimum detectable HR at 80% power, 0.05 alpha, sample size of 201, event rate of 24.88% and variant-specific minor allele frequency.

The two replicated variants and their risk alleles associated with increased mortality risk were rs4526713-T, (additive model), and rs4149117-T, (dominant model). Both the risk alleles and their inheritance models were consistent with original publications. Figure 2A-B,

**Figure 2.**
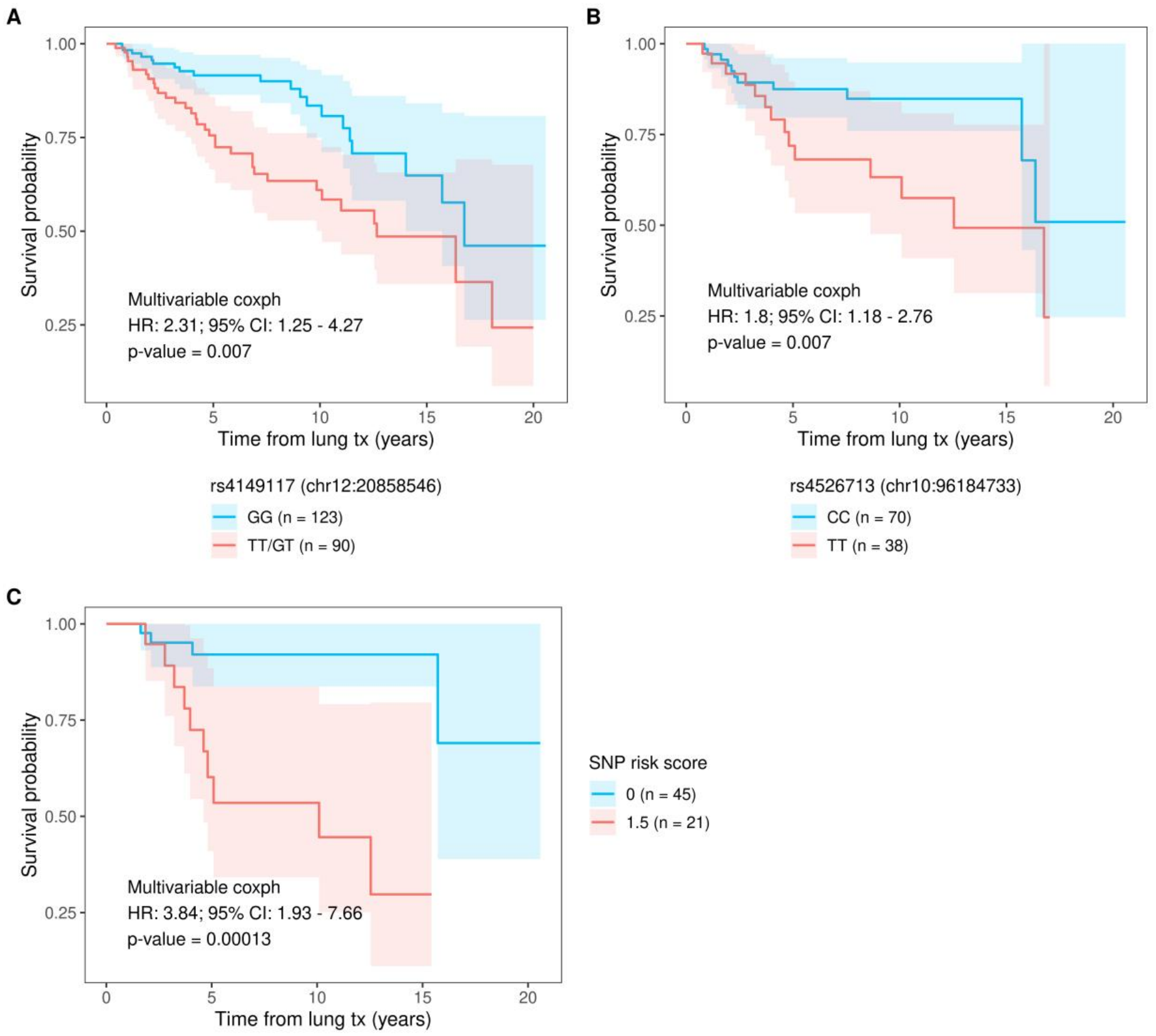
Kaplan-Meier curves and multivariable coxph statistics of top replicated SNPs.A) Dominant effect of rs4149117-T. B) Additive effect of rs4526713-T, showing curves for homozygotes. C) Effect of score constructed from the two top SNPs in A) and B). The multivariable coxph statistics refer to a continuous score while the Kaplan-Meier survival graphs show effect of the top and bottom score quantiles. Shaded areas around the survival curves show 95% confidence intervals.

### Independent effects of replicated variants

We included the replicated SNPs in the same multivariable coxph model to study their independent contribution to survival after LTx. Their associations with survival remained directionally consistent and significant at p < 0.01, supporting independent contributions to post-transplant mortality risk (Table 3, Table S3).

**Table 3.** Multivariable coxph results for independent effects of the replicated SNPs.

| **rsID** | **chr:position_ref:alt** | **Risk genotype** | **Genetic model** | **p-value** | **HR** | **95% CI** |  |
|---|---|---|---|---|---|---|---|
|  |  |  |  |  |  | **Lower** | **Upper** |
| rs4526713 | 10:96184733:C:T | T | additive | 0.0069 | 1.80 | 1.16 | 2.76 |
| rs4149117 | 12:20858546:T:G | T | dominant | 0.0073 | 2.31 | 1.25 | 4.27 |

### Two-SNP genetic risk score

The two-SNP score showed a stronger association with mortality than any individual SNP. Kaplan–Meier visualization suggested separation between patients in the upper and lower tails of the score distribution, with the highest-risk group showing poorer survival than the lowest-risk group (Figure 2C, Table S3).

### Plasma protein QTL

Extraction of plasma protein QTL (pQTL) data for the replicated variants rs4526713 and rs4149117 from the FinnGen-UK Biobank meta-analysis summary statistics available in FinnGen identified three statistically significant associations after Bonferroni correction for the number of SNPs and proteins tested (Table S4). All significant pQTLs were associated with rs4149117, which was linked to the plasma levels of MEGF9, MET, and PTPRF.

## Discussion

In this candidate-variant replication study of 201 Finnish LTx recipients from FinnGen, we identified three independent variants whose associations with post-transplant survival were concordant with prior reports. The strongest signal was observed for an aggregate two-SNP score, suggesting that inherited variants may jointly contribute to survival after lung transplantation. These results support the hypothesis that recipient genetic background modifies post-transplant outcomes, although the findings should be interpreted as hypothesis-strengthening rather than definitive clinical evidence.

The finding involving rs4149117 is biologically and clinically plausible because this variant is located in *SLCO1B3* regulating its activity, a transporter gene previously implicated in outcomes among lung-transplant recipients treated with mycophenolic acid (Tague et al., 2020). In the original pharmacogenomic study, *SLCO1B3* variants rs4149117 and rs7311358 were associated with decreased one-year survival, increased risk of nonminimal acute rejection, and lower survival through three years among mycophenolic-acid-treated lung-transplant recipients. PharmGKB/ClinPGx annotations summarize rs4149117 T-carrier status as associated with reduced overall survival and increased CLAD risk in lung-transplant recipients receiving mycophenolic acid (https://www.clinpgx.org/variantAnnotation/1451102844), although the evidence level remains moderate and not yet practice-changing. Furthermore, rs4149117 risk allele downregulates plasma protein MET, receptor for hepatocyte growth factor (HGF), which promotes epithelial repair, wound closure, cell survival, and anti-fibrotic responses in lung injury models (Atanelishvili et al., 2016). The replication of rs4149117 signal in FinnGen therefore supports further evaluation of HGF and immunosuppression-related pharmacogenomic modifiers of lung-transplant survival.

The other replicated variant, rs4526713, located in *BLNK*, could plausibly influence survival after LTx by modifying immune pathways in B-cell humoral alloimmunity. *BLNK* encodes B-cell linker protein, an adapter molecule in B-cell receptor signaling. B cells are increasingly recognized as active contributors to alloimmune injury in LTx setting, not only through antibody production but also through antigen presentation, tertiary lymphoid-organ formation (Zhao et al., 2024), and local immune amplification inside chronically rejected grafts (Ohm & Jungraithmayr, 2022). This (Chen et al., 2018)(Watanabe et al., 2023)pathway could accelerate infection-triggered inflammation, rejection episodes, or progressive airway fibrosis. This model fits current concepts of CLAD as a complex syndrome involving alloimmune injury, infection or airway inflammation, epithelial damage, vascular and airway remodeling, and fibrosis (Renaud-Picard et al., 2024).

A strength of the present study is its independent population-based setting. FinnGen provides longitudinal health-record linkage and genome-wide genotype data, allowing candidate associations to be evaluated in a Finnish cohort outside the original discovery populations (Kurki et al., 2023). The study also attempted to assess genetic architecture beyond single markers by combining replicated variants into an aggregate score.

However, several limitations are important. First, the sample size was modest for genetic survival analysis, particularly for low-frequency variants and recessive models. Second, the endpoint was all-cause mortality rather than CLAD, graft loss, acute rejection, or cause-specific death. Although death after lung transplantation is clinically meaningful and often linked to graft dysfunction (Porteous & Lee, 2017), it is etiologically heterogeneous. Third, donor genotypes, donor–recipient HLA matching, immunosuppressive protocols, infection history, primary graft dysfunction, rejection episodes, and CLAD phenotype were unavailable. These missing variables may confound or mediate genetic associations. Furthermore, although only two of the evaluated candidate variants were replicated in our analysis, the absence of significant associations for the remaining variants does not exclude a potential role in LTx outcomes. Rather, the limited statistical power of our dataset may have reduced our ability to detect these signals.

Despite these limitations, the results provide useful replication evidence. The strongest immediate implication is not clinical genotyping, but prioritization of replicated variants for larger studies with harmonized transplant phenotypes. Future work should evaluate these SNPs in multicenter cohorts with recipient genotypes, detailed immunosuppression data, CLAD assessment, rejection phenotypes, HLA molecular mismatch, and cause-specific mortality. If replicated further, a multi-variant risk score could eventually contribute to post-transplant risk stratification, pharmacogenomic tailoring, intensified monitoring strategies, and suggest new therapeutic targets.

## Conclusions

This study replicated two previously reported SNP associations with survival after lung transplantation in an independent Finnish cohort. The combined SNP score showed stronger association with mortality than individual variants, suggesting that genetic variation may contribute cumulatively to post-transplant survival. Larger, phenotype-rich donor–recipient genomic studies are needed to validate these findings and determine whether genetic risk profiling can improve clinical management after lung transplantation and provide mechanistic insights.

## Data Availability

All analysis summary statistics data are available as Supplementary Tables. Individual-level participant data from FinnGen cannot be made publicly available by the authors due to data protection restrictions. Access to individual-level sensitive health data is governed by Finnish and European legislation, including the General Data Protection Regulation (GDPR). Researchers may apply for access to Finnish health registry data through Findata (https://findata.fi/en/permits/) and for individual-level genotype data through the Fingenious portal (https://site.fingenious.fi/en/), maintained by the Finnish Biobank Cooperative (FINBB) (https://finbb.fi/en/), subject to the relevant regulatory approvals.

## Acknowledgements

The FinnGen project is funded by two grants from Business Finland (HUS 4685/31/2016 and UH 4386/31/2016) and the following industry partners: AbbVie Inc., Alnylam Pharmaceuticals, Inc., AstraZeneca UK Ltd, Bayer AG, Biogen MA Inc., Boehringer Ingelheim International GmbH, Bristol Myers Squibb Inc. (and Celgene Corporation & Celgene International II Sàrl), Genentech Inc., GlaxoSmithKline Intellectual Property Development Ltd., Johnson&Johnson Innovative Medicine Inc., Maze Therapeutics Inc., Merck Sharp & Dohme LCC, Novartis AG, Pfizer Inc. and Sanofi US Services Inc. Following biobanks are acknowledged for delivering biobank samples to FinnGen: Auria Biobank (www.auria.fi/biopankki), THL Biobank (www.thl.fi/biobank), Helsinki Biobank (www.helsinginbiopankki.fi), Biobank Borealis of Northern Finland (https://www.ppshp.fi/Tutkimus-ja-opetus/Biopankki/Pages/Biobank-Borealis-briefly-in-English.aspx), Finnish Clinical Biobank Tampere (www.tays.fi/en-US/Research_and_development/Finnish_Clinical_Biobank_Tampere), Biobank of Eastern Finland (www.ita-suomenbiopankki.fi/en), Central Finland Biobank (www.ksshp.fi/fi-FI/Potilaalle/Biopankki), Finnish Red Cross Blood Service Biobank (www.veripalvelu.fi/verenluovutus/biopankkitoiminta), Terveystalo Biobank (www.terveystalo.com/fi/Yritystietoa/Terveystalo-Biopankki/Biopankki/) and Arctic Biobank (https://www.oulu.fi/en/university/faculties-and-units/faculty-medicine/northern-finland-birth-cohorts-and-arctic-biobank). All Finnish Biobanks are members of BBMRI.fi infrastructure (https://www.bbmri-eric.eu/national-nodes/finland/). Finnish Biobank Cooperative - FINBB (https://finbb.fi/) is the coordinator of BBMRI-ERIC operations in Finland. The Finnish biobank data can be accessed through the Fingenious^®^ services (https://site.fingenious.fi/en/) managed by FINBB.

